# Consideration for the asymptomatic transmission of COVID-19: Systematic Review and Meta-Analysis

**DOI:** 10.1101/2020.10.06.20207597

**Authors:** Khaiwal Ravindra, Vivek Singh Malik, Bijaya K Padhi, Sonu Goel, Madhu Gupta

## Abstract

**Objective:** Worldwide countries are experiencing viral load in their population, leading to potential infectivity of asymptomatic COVID-19. Current systematic review and meta-analysis aimed to investigate the role of asymptomatic infection worldwide reported in family-cluster, adults, children, health care workers, and travelers.

**Design:** Online literature search (PubMed, Google Scholar, medRixv, and BioRixv) was accomplished using standard Boolean operators, studies published till 07^th^ June 2020.

**Setting:** Studies were included from case reports, short communication, and retrospective to cover sufficient asymptomatic COVID-19 transmission reported.

**Participants:** Familial-clusters, adults, children, health care workers, and travelers.

**Results:** We observed asymptomatic transmission among familial-cluster, adults, children, health care workers, and travelers with a proportion of 32% 37%, 26%, 6%, and 32%, respectively. This study observed an overall proportion of 31% (95%CI: 0.19-0.44) with heterogeneity of I^2^ (97.28%, p=<0.001) among all asymptomatic populations mentioned in this study. Among children and healthcare workers, this study showed no heterogeneity; to overcome the interpretation from a fixed model, the random effect model was also applied to estimate the average distribution across studies included in the meta-analysis.

**Conclusion:** We found and suggest the rigorous epidemiological history, early isolation, social distancing, and increased quarantine period (at least 28 days) after screening asymptomatic cases as well as their close contacts for chest CT scan even after their negative nucleic acid testing to minimize the spread among the community. This systematic review and meta-analysis support asymptomatic COVID-19 transmission between person to person depending on the variation of virus incubation period among individuals. Children especially, school-going aged <18 years, need to be monitored and prevention strategy, e.g., chest CT and social distancing required to prevent the community transmission of COVID-19 in asymptomatic mode.

**Strengths and limitations of this study:**

- Examine the possibility of asymptomatic COVID-19 transmission in the community at different levels.
- Supports contact tracing, social distancing, early isolation, and increased quarantine period to minimize the risk of virus spread.
- Supports chest CT scan and viral nucleic acid testing to identify the asymptomatic cases in the community.
- Supports rigorous epidemiological history with multiple detection methods.
- A higher proportion of asymptomatic incidence was seen, suggests monitoring, and maintaining social distancing.

## Introduction

Symptomatic viral infections have been a significant risk factor for the public. It is more concerned while asymptomatic viral infection occurs in the community. For symptomatic cases, fever, dyspnea, dry cough, and diarrhea are the major sign, and symptoms were reported lasting up to 14 days with a median incubation period 9-12 days. Aerosol transmissions occurred through sneezing or coughing and reported to be the primary route of infection from person to person ^1^. Simulation studies have been done and observed asymptomatic transmission among person-person^2^. PCR-based assays were suggested in managing the asymptomatic transmission of the virus by carriers ^3^. The first case of asymptomatic transmission of COVID-19 was reported by JAMA on 21^st^ February 2020 by an asymptomatic carrier (He et al., 2020; Wang et al., 2020). Asymptomatic infection was reported as “hidden coronavirus infections” (“infections” or “covert coronavirus infections” ^5^. Asymptomatic COVID-19 cases should be quarantined for 14 days, and their nucleic acid test should be negative twice before discharging, which is mentioned in the COVID-19 prevention and control protocol (6^th^ edition). Worldwide, interest in asymptomatic COVID-19 infections and their transmission potential has been increased ^6^. In China, around 86% asymptomatic COVID-19 transmission was undocumented before travel restrictions ^6^.

Till now, asymptomatic COVID-19 cases have been reported among Familial-cluster ^7–12^, pregnant women ^13,14^, adults ^15–24^, children ^1,25,26^, health care worker ^27–29^, and travelers ^30–34^. Considering the potential transmission of asymptomatic COVID-19 among the community we tried to accumulate the desired information from the general population as well as vulnerable groups from the different backgrounds were taken, and meta-analysis was performed. There are no previous studies available for asymptomatic COVID-19 transmission among different subgroups between person-person.

## Methods

### Search Strategy

For the meta-analysis PRISMA guideline was applied on this study ^35,36^, Boolean operators “asymptomatic transmission”, “((COVID-19) AND (Coronavirus)) AND (Asymptomatic transmission)”, “((COVID-19) OR (Coronavirus)) AND (asymptomatic transmission)”, “(SARS-CoV-2) AND (asymptomatic transmission)”, “(2019-nCoV) AND (asymptomatic transmission)”, “(Wuhan pneumonia) AND (asymptomatic transmission)”, “(Wuhan flu) AND (asymptomatic transmission)”, “(2019-nCoV acute respiratory disease) AND (asymptomatic transmission)”, “(2019-nCoV respiratory syndrome) AND (asymptomatic transmission)” used for the PubMed database, Google scholar, medRxiv, and BioRixv. The study included published literature without language restriction until 07^th^ June 2020. Selection criteria (Inclusion/exclusion criteria): studies with the following conditions were included for the meta-analysis: (1) Case reports, case series, and cohort study, (2) Asymptomatic infection of COVID-19 (clinical, laboratory or both confirmed), and (3) Studies reporting cross-sectional were excluded.

### Data extraction

Details of authors, sample size, and numbers reported for the asymptomatic infection of COVID-19 were extracted and recorded independently. Data extraction was done separately by two independent reviewers and disagreement was settled by joint discussion. To minimize the risk of duplication of data was carefully handled.

### Quality assessment

The Newcastle Ottawa scale (cohort studies) was used to evaluate the selected studies in the current systematic review and meta-analysis ^35,37^.

### Publication bias

Possible publication bias was not calculated in this study as we have included cohort, case report study design to cover the possible asymptomatic cases considering the current situation causing limited power of the among studies ^38^.

### Statistical analysis and data synthesis

After extracting the results, studies were pooled, and the effect of asymptomatic transmission of COVID-19 was examined through the random effects method. For continuous outcome standard error (SE) with 95% CI was calculated. The heterogeneity (*I*^2^statistic) was assessed between studies. *I*^2^ values the existence of heterogeneity was taken, as suggested by Higgins and colleagues ^35,39,40^. Data for meta-analysis was accomplished as described by ^35,41^.

### Patient and public involvement

There was no direct patient or public involvement in this systematic review and meta-analysis.

### Results: Literature search

Literature search and screening were done according to the PRISMA chart (Figure: 1). Initially, 4,667 published research articles were identified using online database search. After the removal of 4617 Publications due to duplications, only 50 research articles were shortlisted, followed by further screened for relevance. Finally, 27 articles meeting the inclusion criteria were grouped into the family-cluster (total studies: 06), Adults (total studies: 10), Children (total studies: 03), health care workers (total studies: 03), and travelers (total studies: 05) were included in the quantitative synthesis of the current study.

**Figure 1:**
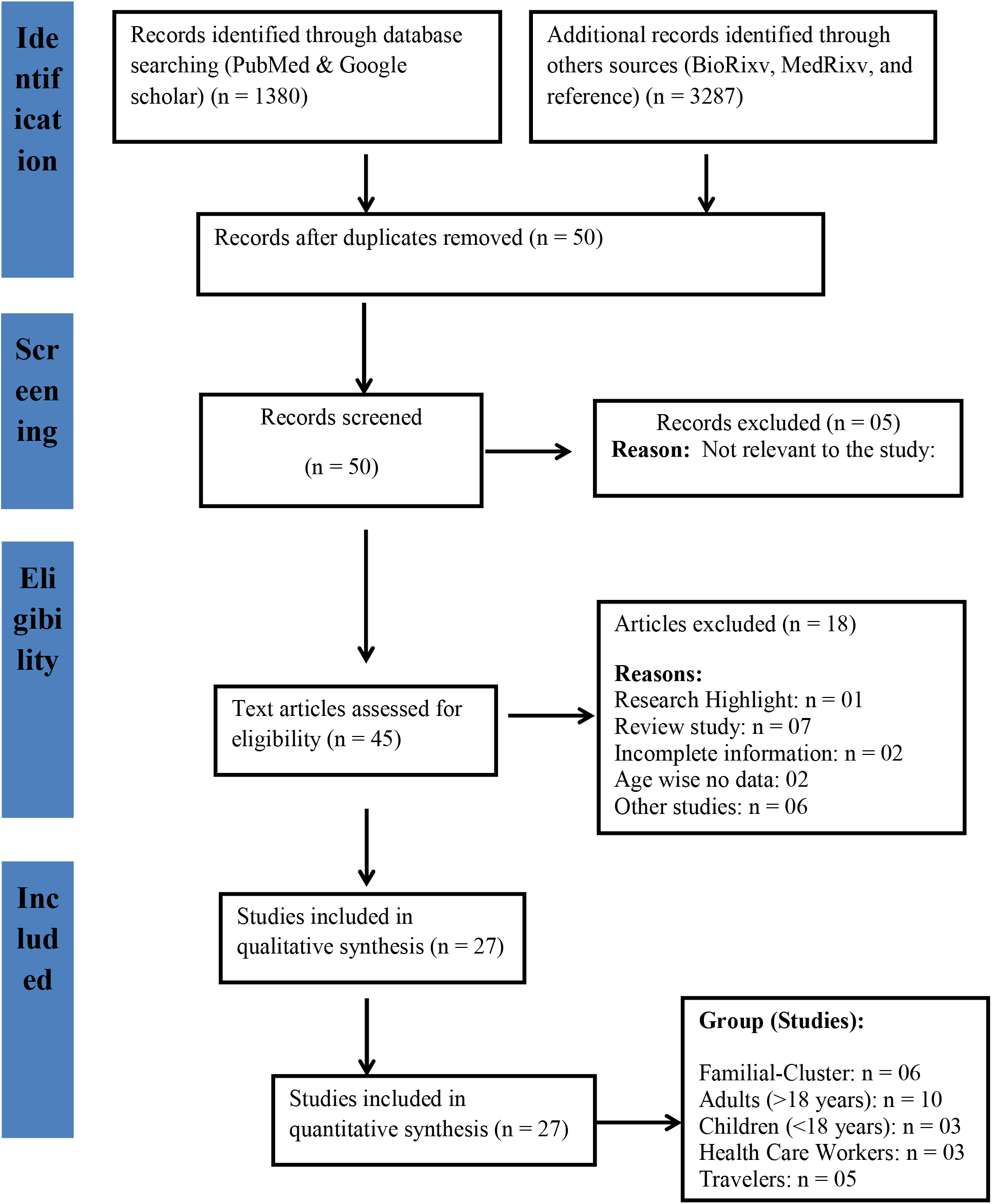
PRISMA Chart.

### Characteristics of the Study

The main components of the studies are summarized (Table 1). All published research articles fall under the cohort (observational) study design. Most of the studies are from China, Korea, USA, Japan, and Germany. The study included articles published/available online till 07^th^ June 2020.

**Table 1:** Characteristics of the study participants.

| Author | Country | Age, years<br>(mean (±SD)/<br>median(IQR)) | Study Type | Type of<br>test | Major findings |
| --- | --- | --- | --- | --- | --- |
| **Family cluster** |  |  |  |  |  |
| Jasper Fuk-woochan | China | Family: 36-60<br>Child: 10 | Cohort | RT-PCR | Supports person to person transmission between family |
| Ming Chen | China | 8.5 ± 0.17 | Case report | RT-PCR | The ability of COVID-19 transmission during the asymptomatic period even after negative viral testing |
| Shubia Lu | China | 8 | Case report | RT-PCR | Supports rigorous investigation in the combination of various testing methods to for asymptomatic COVID-19 cases |
| GuoaingQian | China | 6 | Brief report | RT-PCR | Variation in clinical manifestation across individuals was observed |
| Feng Ye | China | 38 ± 18.38 | Cohort | RT-PCR | Possibility of COVID-19 transmission by the asymptomatic carrier during the incubation period |
| Yan Bai | China | 20 | Cohort | RT-PCR | Support asymptomatic transmission through a family contact |
| **Adults** |  |  |  |  |  |
| SijiaTian | China | 47.5 | Cohort | RT-PCR | Early isolation and quarantine for close contacts to prevent asymptomatic transmission |
| G.-u. Kim | Korea | 26 (22-47) | Research note | RT-PCR | Supports social distancing to prevent asymptomatic transmission |
| Wei Fang Kong | China | 37.7 ($\pm 19$) | Cohort | RT-PCR | Suggest rigorous epidemiological history and chest CT scan as a practical tools to identify the asymptomatic COVID-19 cases in the community |
| Guosheng Yin | China | - | Cohort | RT-PCR | No difference in the transmission rate of COVID-19 between asymptomatic and symptomatic cases |
| HengMeng | China | 42.60 ($\pm 16.56$) | Cohort | RT-PCR | Suggest chest CT scan as an important tool to screen the asymptomatic COVID-19 cases in the community |
| Farida Ismail AI Hosani | UAE | 37 (30-45) | Cohort | RT-PCR | No transmission among household contacts after the implication of strong isolation policies |
| Daihie He | China | - | Cohort | RT-PCR | Significantly smaller transmissibility of asymptomatic cases than symptomatic |
| ChengfengQiu | China | 43 (8-84) | Cohort | RT-PCR | Transmission occurred after 14 days quarantine periods, suggests |
| Rui Zhou | China | - | Short communication | RT-PCR | Suggest rigorous epidemiological history and nucleic acid testing |
| Shin Young Park | Korea | 38 (20-0) | Report | RT-PCR | Supports contact tracing, testing and increasing quarantine to prevent asymptomatic COVID-19 transmission in the community |
| **Children** |  |  |  |  |  |
| Zhiliang Hu | China | <15 | Cohort | RT-PCR | Suggest close contact tracing, and nucleic acid testing to identify the asymptomatic COVID-19 tracing the community |
| HaiyanQiu | China | 8.3 ($\pm 3.5$) | Cohort | RT-PCR | Possibility of asymptomatic COVID-19 transmission by close contacts |
| Xin Tan | China | - | Cohort | RT-PCR | Possibility of asymptomatic COVID-19 transmission by intra familial contact |
| **Health Care Workers** |  |  |  |  |  |
| Anne Kimball | USA | - | - | RT-PCR | Reported rapid transmission among health care worker |
| Vera Schoierzeck | Germany | 48 | Cohort | RT-PCR | Suggest nucleic acid testing for asymptomatic COVID-19 cases |
| Jose Lucar | USA | >18 | Cohort | RT-PCR | transmission reported because of prolonged surgery done on asymptomatic COVID-19 case |
| **Traveler>18** |  |  |  |  |  |
| COVID-19 Team, Korea | Korea | >18 | Cohort | RT-PCR | Supports asymptomatic transmission with minor symptoms |
| Kenji Mizumoto | Japan | >18 | Rapid communication | RT-PCR | Support social distancing to prevent the asymptomatic transmission |
| Ren Wan | China | >18 | Short communication | RT-PCR | Possibility of asymptomatic transmission after 14 days quarantine from asymptomatic COVID-19 case |
| Justin WONG | Brunei | - | Rapid communication | RT-PCR | Support social distancing & nucleic acid testing of asymptomatic COVID-19 case |
| ThiQuynh Mai Le | China | - | Abstract | - | Support asymptomatic COVID-19 viral transmissibility in the absence of signs and symptoms |

**Note: -: Missing values (mean/median values were not reported)**

### Quality assessment

The Newcastle Ottawa Scale (for cohort studies) was used for qualitative evaluation of the studies included in the meta-analysis ^35,37^. The risk of bias was assessed based on three domains (selection, comparability, and outcome), as highlighted (Table 2).

**Table 2:**
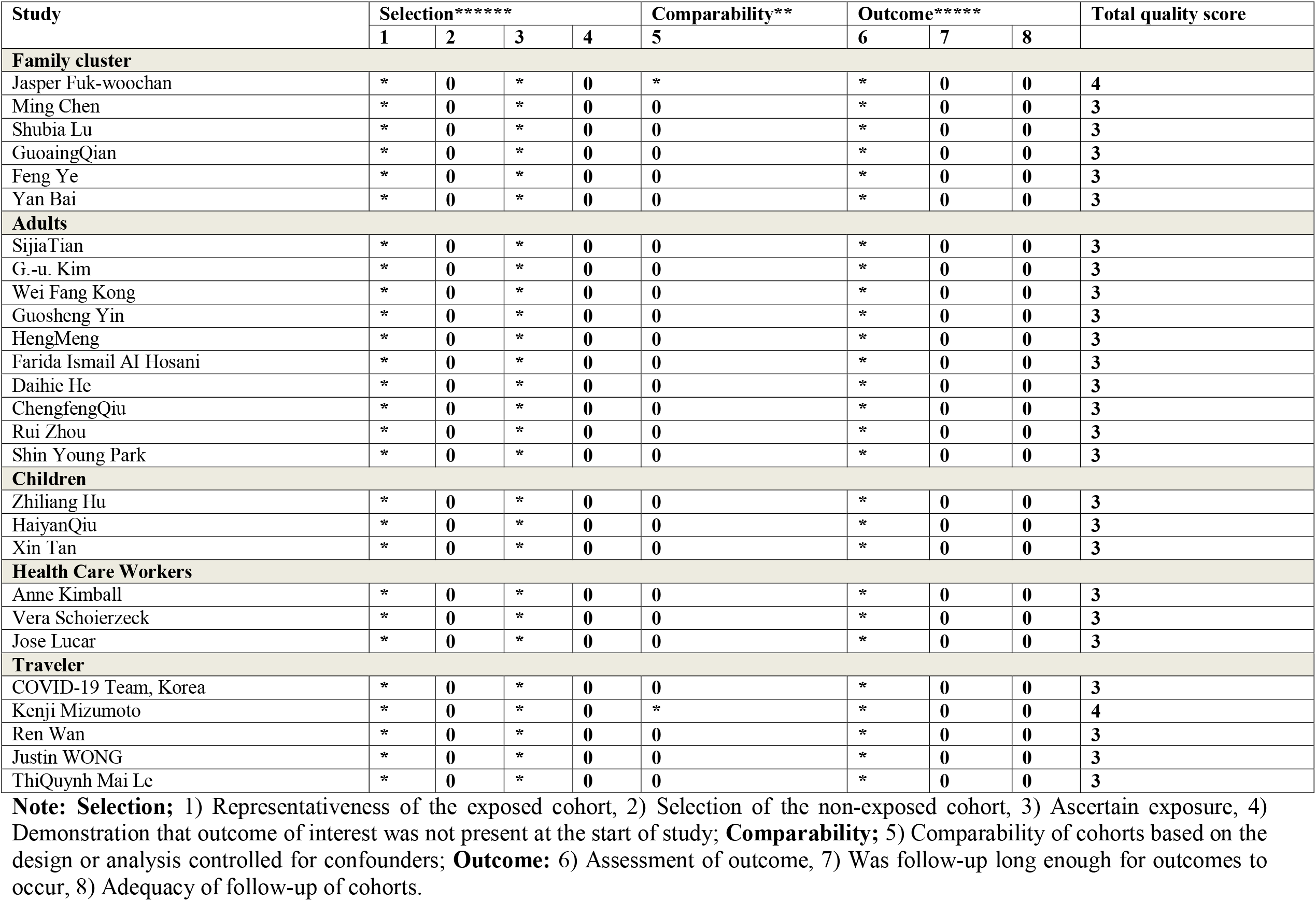
Quality assessment: Cohort study quality according to the Newcastle-Ottawa scale.

### Publication bias

This study included studies published as well as unpublished literature on MedRxic and BioRiv, as long as it meets study inclusion criteria.

### Meta-analysis

The outcomes of the current meta-analysis (Table 3) and forest plot of asymptomatic positivity of COVID-19 among population (Figure 2) and different sub-groups (Figure 3) are shown. A random-effects model was used for the reported asymptomatic COVID-19 transmission at different levels in the community. Current meta-analysis observed heterogeneity among familial-clusters with I^2^= 46.02%, *p*=0.0.10 with proportion of 32%(95%CI= (0.09-0.59), adults (aged >18 years) I^2^=98.44%, p=<0.001 with proportion of 37% (95%CI: 0.19-0.57), Travelers I^2^= 98.18%, *p* = <0.001 with proportion of 32% (95%CI = 0.06-0.65), this study did not observed any heterogeneity among children and health care workers but the proportion of asymptomatic transmission was seen with rate of 26% (95%CI: 0.16-0.37) and 6% (95%CI: 0.00 – 0.31) respectively with overall heterogeneity of I^2^ = 97.28%, p=<0.001. We didn’t observe any significant difference of heterogeneity between groups (p=0.538). With no heterogeneity among children and health care workers random effect model observed an overall proportion of 26% and 6%, respectively. The likelihood ratio (LR) test was applied for random effect vs. fixed effect model to check the weighted difference (Figure 4). This study observed a proportion of 25% (95%CI: 0.17 – 0.35), chi-square value 129.9, p=<0.0001 of asymptomatic positivity of COVID-19 among the population.

**Table 3:** The meta-analysis of asymptomatic transmission for COVID-19 among different sub-groups of the population:

| Risk factor<br>(Asymptomatic) Group | Studies | Asymptomatic<br>Population (n) | Total<br>Sample<br>Size (N) | Proportion<br>(95%CI) | $I^2$ |
| --- | --- | --- | --- | --- | --- |
| Family Cluster | 6 | 9 | 31 | 0.32 (0.09-0.59) | 46.02%, $p = 0.10$ |
| Adults (>18 years age) | 10 | 374 | 1688 | 0.37 (0.19-0.57) | 98.44%, $p < 0.001$ |
| Children (<18 years age) | 3 | 36 | 73 | 0.26 (0.16-0.37) | - |
| Health Care Workers | 3 | 13 | 125 | 0.06 (0.00-0.31) | - |
| Travelers | 5 | 368 | 876 | 0.32 (0.06-0.65) | 98.18%, $p < 0.001$ |
| Combined (groups) | 27 | 813 | 2793 | 0.31 (0.19-0.44) | 97.28%, $p < 0.001$ |

**Figure 2:**
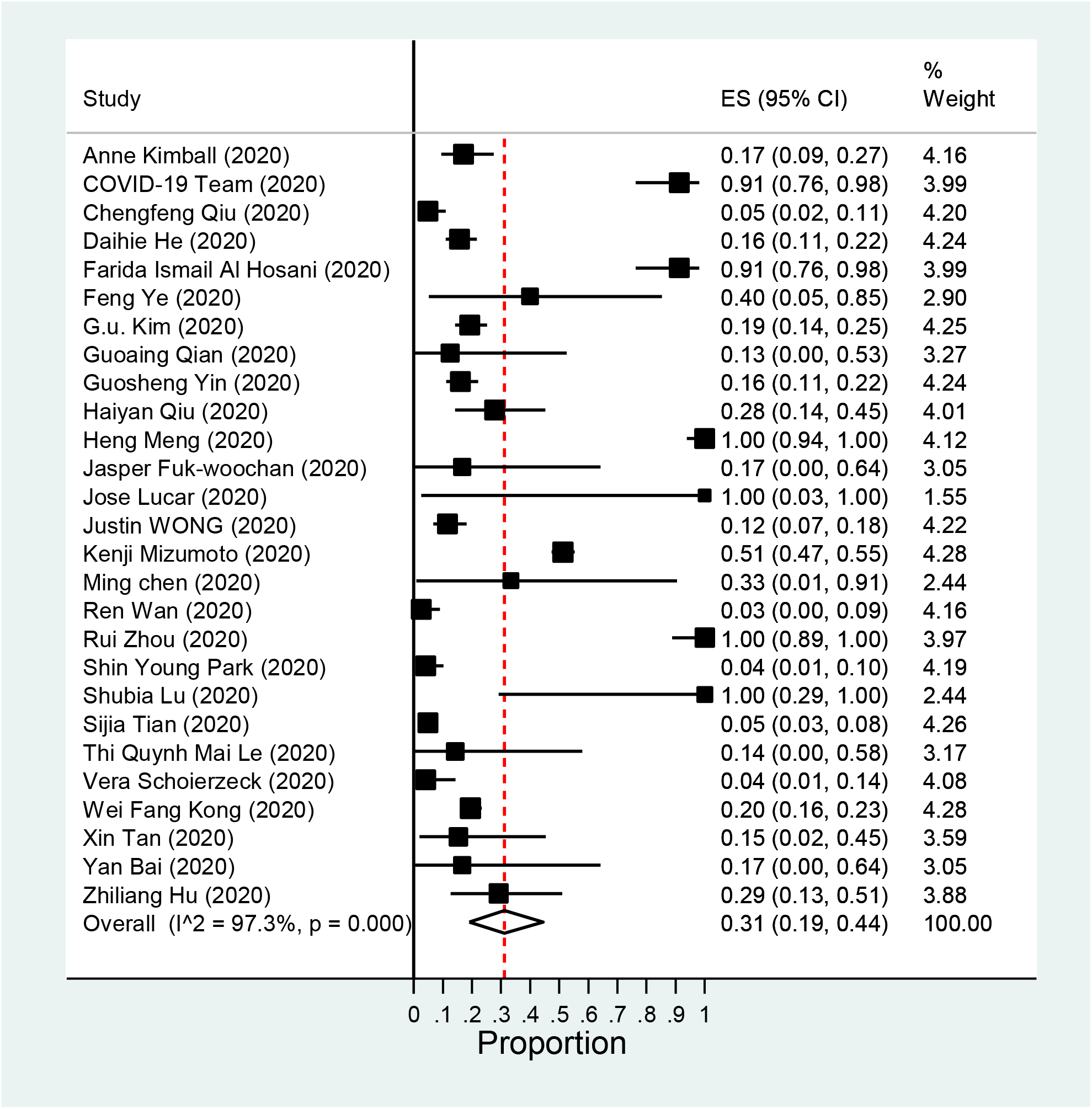
Forest plot asymptomatic positivity of COVID-19 among population.

**Figure 3:**
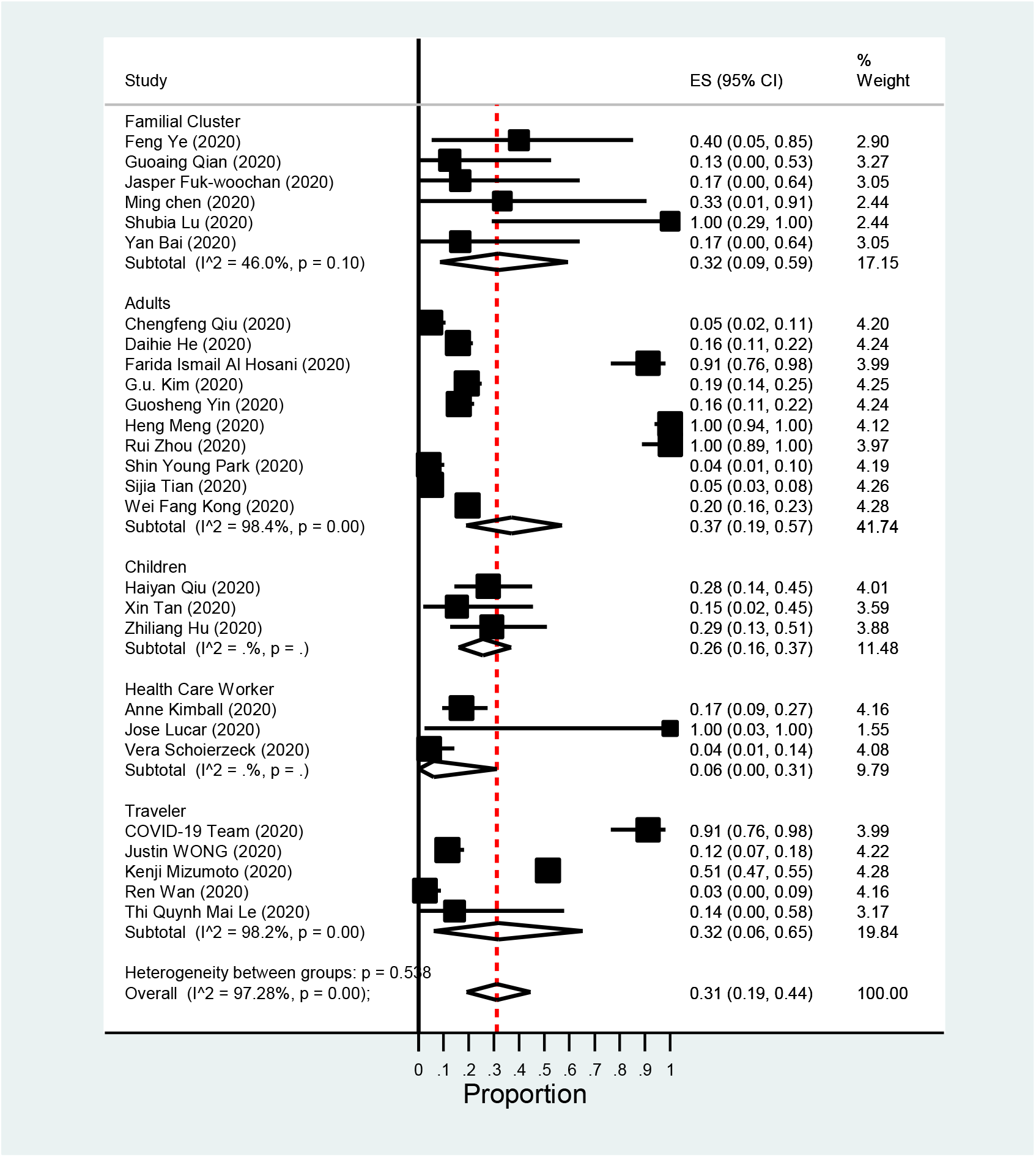
Forest plot asymptomatic positivity of COVID-19 among different sub-groups.

**Figure 4:**
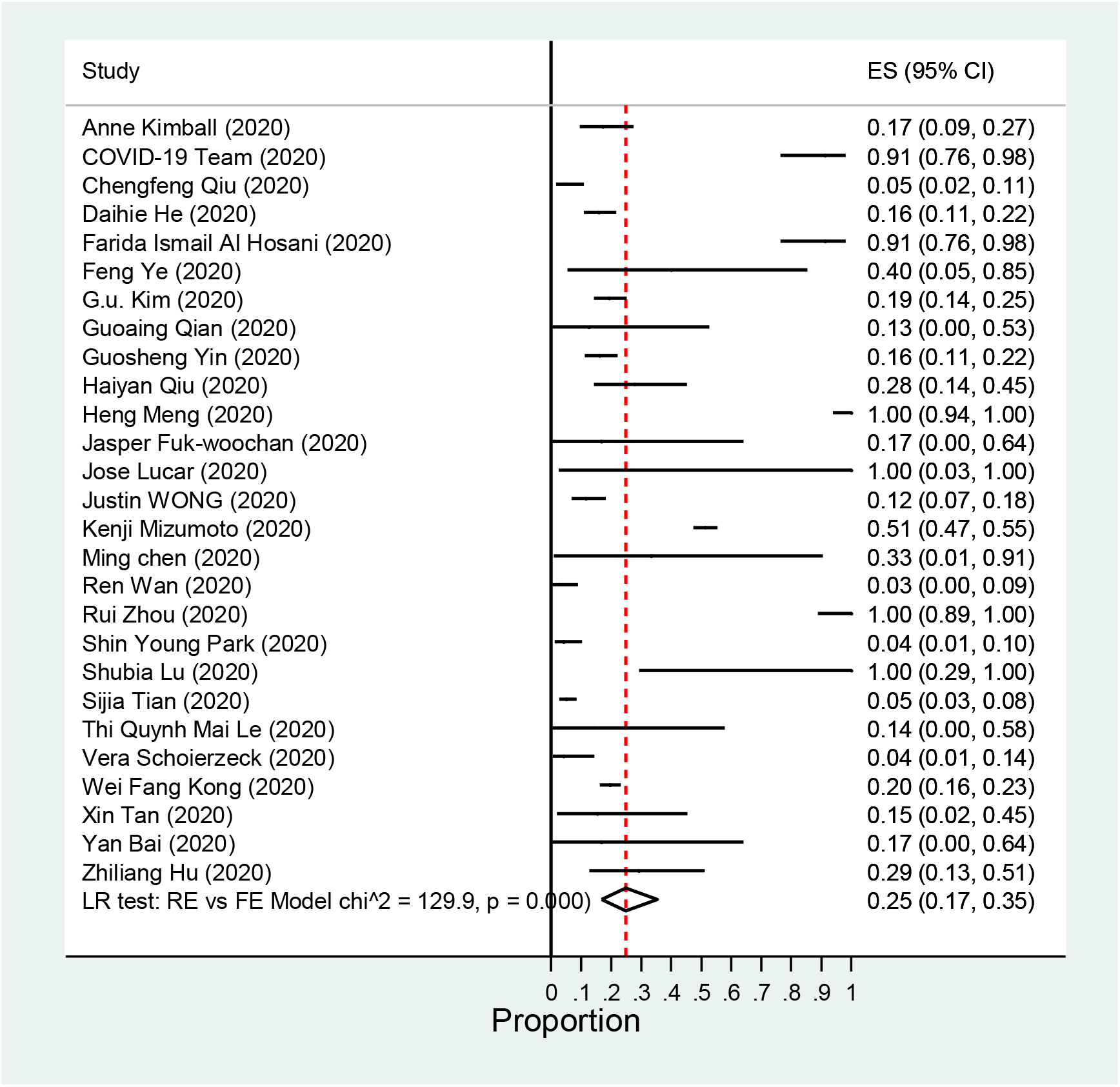
Forest plot: Random Effect and Fixed Effect of Asymptomatic positivity of COVID-19 among population.

## Discussion

The current study summarized available retrospective studies, case reports from family-cluster, adults, children, health care workers, and travelers. Person to person asymptomatic transmission was observed among familial-cluster in an asymptomatic COVID-19 child aged ten years showed abnormal chest CT and another child with mild chest CT manifestation when his family members were diagnosed COVID-19 positive showing sign of fever and respiratory issues ^8,9^. The study suggests that thorough epidemiological investigations in combination with multiple detection methods (e.g., RT-PCR, chest CT, Rapid IgM-IgG, and serum CRP level) can identify the asymptomatic carriers ^10,11^ in the community among varying clinical manifestations between individuals. Another study supports the possibility of asymptomatic transmission among familial-cluster during the incubation period ^12^. In a familial-cluster of 5 positive COVID-19 patients had contact with other asymptomatic family members who returned from the Wuhan, suggestive of asymptomatic transmission ^7^.

Studies included in the current meta-analysis were checked for the likelihood ratio (LR) between random effect and fixed-effect models for the distribution of asymptomatic COVID-19 transmission among the community.

Among pregnant woman’s fetuses are at high-risk during any disease outbreak, in pregnant woman with asymptomatic COVID-19 viral infection delivered a baby negative for the COVID-19 nucleic acid test Suggestive of no vertical transmission among neonates born to COVID-19 infected mothers ^14,42–49^. In Wuhan, a lower fatality rate and higher discharge rate were observed than in Beijing. It is crucial to identify and take necessary control measures among adults of asymptomatic cases to prevent transmission ^21^. In South Korea, 41 asymptomatic adults were identified and were confirmed by RT-PCR out of 213 cases ^17^. Among 100 asymptomatic cases, 60% developed delayed symptoms, and none of the asymptomatic cases died, suggesting asymptomatic transmission during the incubation period ^18^. Another study did not observe any difference in the symptomatic and asymptomatic COVID-19 transmission rates among patients^22^. In adults, CT imaging of asymptomatic COVID-19 individuals has advantages in highly suspicious cases with negative nucleic acid testing ^17^. In adults, a serological investigation among 31 out of 34 cases with asymptomatic infection did not require oxygen support during hospitalization ^15^. Theoretically, the quantified infection transmission rate shows the estimated risk ratio (RR) of infectivity [3.9%, (95%CI: 1.5 – 11.8)] of symptomatic against asymptomatic.

In asymptomatic adults, the transmission was significantly smaller than that of the symptomatic cases ^16^. No gender difference among males and females was observed for asymptomatic transmission ^20^.

Further longitudinal surveillance via virus nucleic acid testing is warranted to identify and control the viral load among asymptomatic CVOID-19 cases ^24^ in adults. In a study, 4 asymptomatic cases were quarantined for 14 days, and none was able to transmit the infections due to managing proper isolation and quarantine of the cases ^19^.

Asymptomatic COVID-19 transmission was seen in children ^26^. In a study, 24 asymptomatic cases were screened from close contacts of asymptomatic COIVD-19 ^25^. Another study supports the multiple site sampling of close contacts should be performed ^1^ among children. In a review, it was observed that the adults as compared to children with COVID-19 infection show mild clinical symptoms and radiological manifestations as previously reported for SARS- and the Middle East respiratory syndrome (MERS)-CoV ^50^.

Health care workers in a nursing facility: Rapid transmission of COVID-19 had been reported in 76 residents cases. and 23 (30.3%) had positive test results, and approximately 13 were asymptomatic on the day of testing, suggesting the possibility of asymptomatic transmission of COVID-19 ^27^. Establishing effective infection control strategies to prevent the transmission among frontline health care workers and patients from COVID-19 infections should be managed urgently and on priority. In another study, out of 48 participants observed, two asymptomatic cases become positive, suggesting appropriate symptom-based testing strategies are essential to prevent outbreaks of COVID-19 within hospital settings ^29^. In the USA, health care workers not wearing respirators were exposed to an asymptomatic CVOID-19 patient without developing clinical illness ^28^.

Traveler: In Korea, COVID-19 was transmitted by 16 infected travelers from other countries. The disease was infectious at this stage, which resulted from close contacts of asymptomatic carriers ^30^. Most of the infections on board the Diamond Princess Cruise ship highlight the asymptomatic transmission of COVID-19 in confined settings. To further mitigate the transmissibility of COVID-19, it may be advised to minimize the number of gathering in confined settings ^32^. A 36-year-old traveler returned from Wuhan was found COVID-19 positive, and other health workers who were in close contact with the patients were also tested RT-PCR positive. However, the patient initially had no symptoms ^33^. A higher proportion of asymptomatic (12%) was reported among travelers returning to Brunei. In another study, it was suggested to increase the testing facility for asymptomatic COVID-19 cases ^34^. Similarly, an asymptomatic COVID-19 patient showed viral transmissibility without showing any signs and symptoms in travelers from China ^31^. Although, we recommend early isolation for and social distancing for asymptomatic COVID-19 cases, which may led to psychological and emotional loss as reported in qualitative study from UK^51^.

### Limitations

There are some limitations in the current systematic review and meta-analysis. A mixed population, a continuous variable, variation in clinical conditions, and statistical methods may differ and cause heterogeneity among studies included in the current meta-analysis. Further, the study included only asymptomatic COVID-19 transmission reported cases that were included.

### Study Importance

This is the first study reviewing the possibility of asymptomatic COVID-19 transmission at different levels in the community and identified the potential role of isolation, identification of close contacts, social distancing and testing asymptomatic COVID-19 cases with chest CT scan and nucleic acid testing to minimize the spread of the virus in the community.

### The implication of our study

This is the first meta-analysis on the possibility of asymptomatic COVID-19 transmission covering different levels in the community.

## Conclusion

Currently, there is no evidence of COVID-19 transmission ability in the asymptomatic stage, but evidence suggests that asymptomatic infections were not limited from neonates to children and adults. In young people, strong immune status was supposed to be protected against the COVID-19 severity. We hypothesize that the asymptomatic carriers, either children or adults, should be vigilant as they are capable of shielding and transmitting the infection in their incubation period without showing any signs and symptoms. As the evidence supports the involvement of lung function in asymptomatic COVID-19 cases, we recommend the chest CT scans among asymptomatic cases, a useful tool to monitor and trace cases in their incubation period.

## Data Availability

No additional data available.

## Contributors

Dr. Khaiwal Ravindra: Concept design, data extraction, interpretation, final correction and writing first draft. Mr. Vivek Singh Malik: Data extraction, interpretation and writing first draft. Dr. Bijaya K Padhi: Interpretation, internal review of data, review and editing. Dr. Sonu Goel: Discussion, review and editing. Dr. Madhu Gupta: Internal review of data, review and editing.

## Acknowledgment

We thank the Department of Community Medicine and School of Public Health,PGIMER, and Indian Council of Medical Research, New Delhi.

## Pateint consent

not required.

## Data sharing statement

No addditonal data avialable.

